# Detection of SARS-CoV-2 in ascitic fluid: a case of viral peritonitis?

**DOI:** 10.1101/2020.06.25.20139998

**Authors:** Victor C. Passarelli, Ana H. Perosa, Luciano Luna, Danielle D. Conte, Oliver A. Nascimento, Jaquelina Ota-Arakaki, Nancy Bellei

**Affiliations:** Infectious Diseases Division. Department of Medicine. Federal University of São Paulo, São Paulo, Brazil; Pulmonology Division. Department of Medicine. Federal University of São Paulo, Brazil

**Author notes:** **Corresponding author**: Victor C Passarelli, Address for correspondence: Pedro de Toledo Street, 781. Division of Infectious Diseases, Federal University of São Paulo, São Paulo, Brazil.

**Keywords:** coronavirus, ascites, cirrhotic, viral peritonitis, case report

## Abstract

SARS Coronavirus-2 detection in different clinical specimens, including serum and stool, has raised important insights about its dynamics and pathogenesis, but some details remain to be understood.

In that respect, disrupt viral control seen in immunocompromising conditions can help unveil pathogenetic mechanisms and characterize new Coronavirus Disease-19 immunological and clinical aspects in general.

We herein report a case of cirrhosis decompensation with ascites due to COVID-19 and unprecedented detection of SARS-CoV-2 in ascitic fluid, hence a possible acute viral peritonitis, as well as in serum, leading to a fatal disseminated infection.

## Introduction

SARS Coronavirus-2 affects primarily upper and lower respiratory tract, but detection in different clinical specimens has raised important insights about its kinetics and pathogenesis^1^. However, virus’ replication in other body sites and pathogenetic details still remain to be understood^2^, although endothelial injury with intracellular viruses as well as immune system dysregulation have been described^3,4^.

In that respect, disrupt viral control seen in immunocompromising conditions can help unfold new pathogenetic mechanisms and characterize new Coronavirus Disease-19 immunological and clinical aspects.

We herein report an unprecedented case of cirrhosis decompensation with ascites due to COVID-19 and SARS-CoV-2 detection in peritoneal fluid, as well as in serum, leading to a fatal disseminated infection in a kidney transplant patient.

### Case report

A 75-year-old white male with a history of hypertension and a kidney transplantation in 2017 presented in the emergency department on May 18th, 2020, five days following the onset of dry cough and progressive dyspnea, without fever or other respiratory or gastrointestinal tract symptoms. He was on immunossupressive therapy with 4 mg tacrolimus and 5 mg prednisone. Physical examination at admission revealed an axillary temperature of 37.1°C, blood pressure of 160/90 mmHg, heart rate of 104 per minute, respiratory rate of 25 breaths per minute and oxygen saturation of 92% while breathing ambient air. Abdominal examination demonstrated mild ascites with collateral circulation.

He was diagnosed in 2018 with liver cirrhosis, portal hypertension and thrombosis, without a conclusive etiology, but denied any prior decompensation. His previous serologies were negative for HIV, Hepatitis C, Syphilis, Schistosomiasis and he had an spontaneous cure of Hepatitis B with undetectable viral load.

Initial laboratory findings showed normal white cell count but severe lymphopenia (162 per mm^3^) as well as other cirrhosis dysfunctions such as thrombocytopenia, increased international normalized ratio (INR) and hypoalbuminemia. Creatinine was 1.5 times its baseline value (2.4 mg/dl), procalcitonin was elevated at 3.92 ng/dl and d-dimer was above 20 µg/mL (maximum test detection). Additional laboratory exams may be seen on table 1.

**Table 1.**
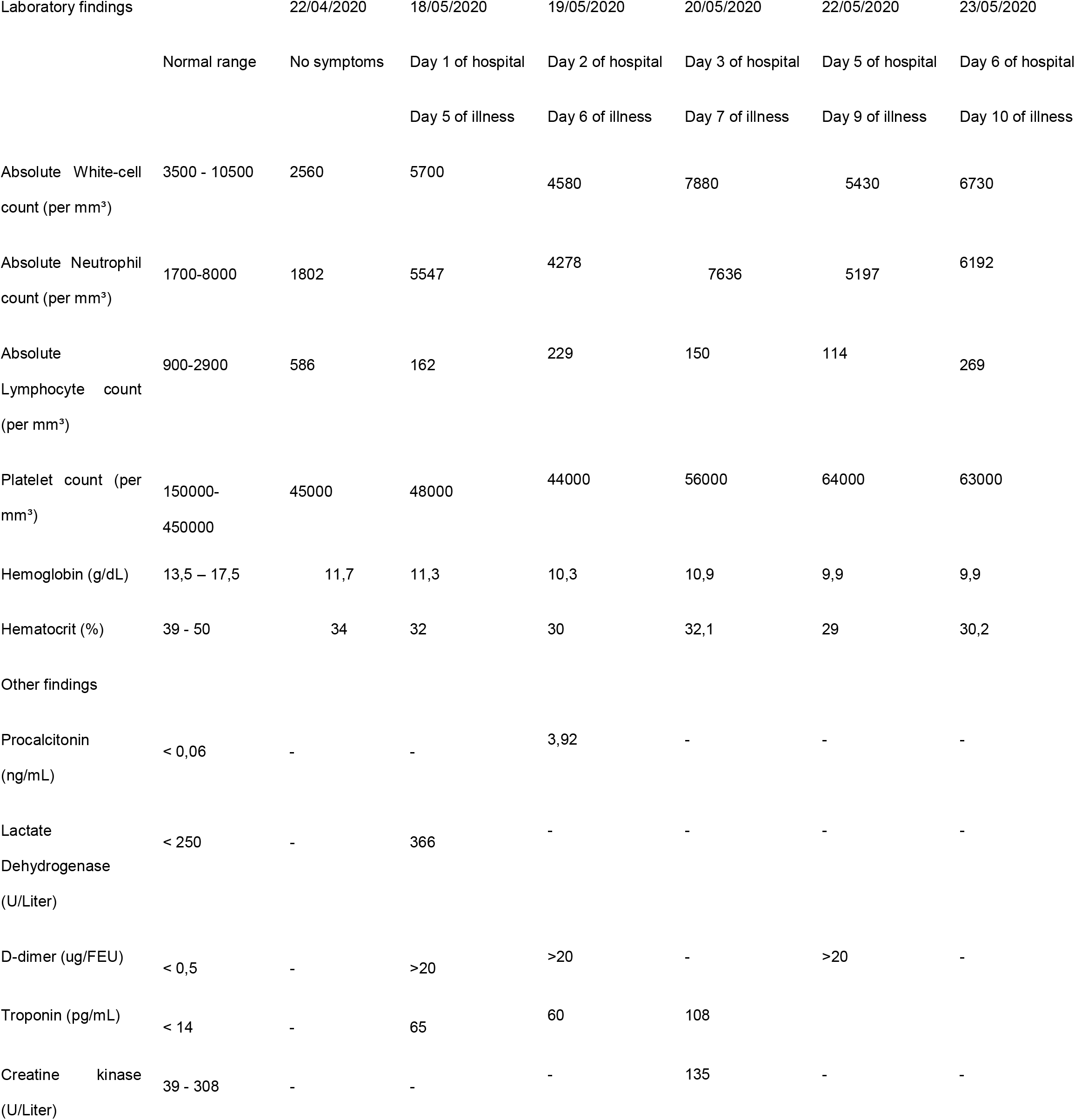
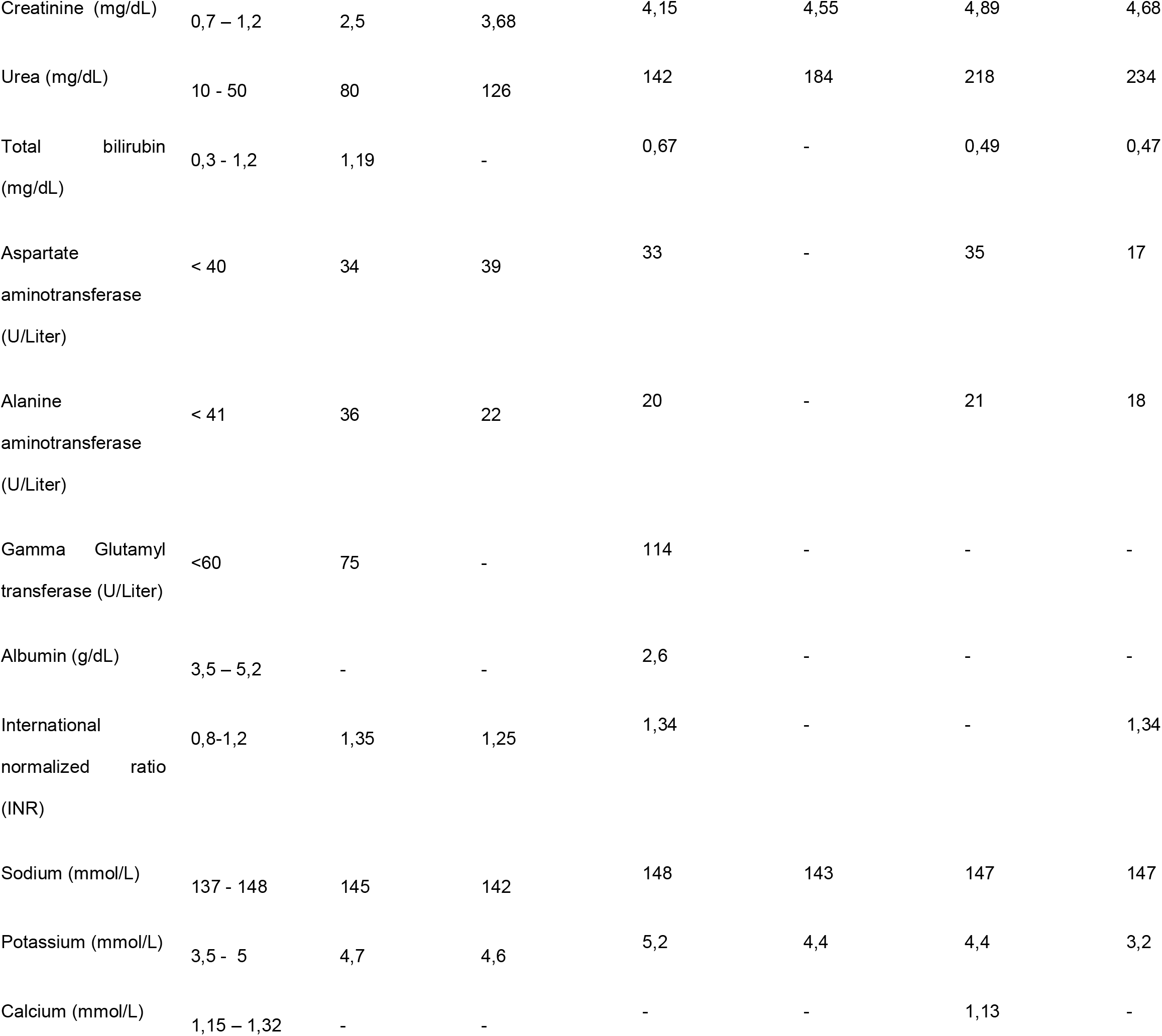
Evolutive blood test results on initial days of hospitalization compared with previous results.

| Laboratory findings |  | 22/04/2020 | 18/05/2020 | 19/05/2020 | 20/05/2020 | 22/05/2020 | 23/05/2020 |
| --- | --- | --- | --- | --- | --- | --- | --- |
|  | Normal range | No symptoms | Day 1 of hospital | Day 2 of hospital | Day 3 of hospital | Day 5 of hospital | Day 6 of hospital |
|  |  |  | Day 5 of illness | Day 6 of illness | Day 7 of illness | Day 9 of illness | Day 10 of illness |
| Absolute White-cell count (per mm<sup>3</sup>) | 3500 - 10500 | 2560 | 5700 | 4580 | 7880 | 5430 | 6730 |
| Absolute Neutrophil count (per mm<sup>3</sup>) | 1700-8000 | 1802 | 5547 | 4278 | 7636 | 5197 | 6192 |
| Absolute Lymphocyte count (per mm<sup>3</sup>) | 900-2900 | 586 | 162 | 229 | 150 | 114 | 269 |
| Platelet count (per mm<sup>3</sup>) | 150000-450000 | 45000 | 48000 | 44000 | 56000 | 64000 | 63000 |
| Hemoglobin (g/dL) | 13,5 – 17,5 | 11,7 | 11,3 | 10,3 | 10,9 | 9,9 | 9,9 |
| Hematocrit (%) | 39 - 50 | 34 | 32 | 30 | 32,1 | 29 | 30,2 |
| Other findings |  |  |  |  |  |  |  |
| Procalcitonin (ng/mL) | < 0,06 | - | - | 3,92 | - | - | - |
| Lactate Dehydrogenase (U/Liter) | < 250 | - | 366 | - | - | - | - |
| D-dimer (ug/FEU) | < 0,5 | - | >20 | >20 | - | >20 | - |
| Troponin (pg/mL) | < 14 | - | 65 | 60 | 108 |  |  |
| Creatine kinase (U/Liter) | 39 - 308 | - | - | - | 135 | - | - |
| Creatinine (mg/dL) | 0,7 – 1,2 | 2,5 | 3,68 | 4,15 | 4,55 | 4,89 | 4,68 |
| Urea (mg/dL) | 10 - 50 | 80 | 126 | 142 | 184 | 218 | 234 |
| Total bilirubin (mg/dL) | 0,3 - 1,2 | 1,19 | - | 0,67 | - | 0,49 | 0,47 |
| Aspartate aminotransferase (U/Liter) | < 40 | 34 | 39 | 33 | - | 35 | 17 |
| Alanine aminotransferase (U/Liter) | < 41 | 36 | 22 | 20 | - | 21 | 18 |
| Gamma Glutamyl transferase (U/Liter) | <60 | 75 | - | 114 | - | - | - |
| Albumin (g/dL) | 3,5 – 5,2 | - | - | 2,6 | - | - | - |
| International normalized ratio (INR) | 0,8-1,2 | 1,35 | 1,25 | 1,34 | - | - | 1,34 |
| Sodium (mmol/L) | 137 - 148 | 145 | 142 | 148 | 143 | 147 | 147 |
| Potassium (mmol/L) | 3,5 - 5 | 4,7 | 4,6 | 5,2 | 4,4 | 4,4 | 3,2 |
| Calcium (mmol/L) | 1,15 – 1,32 | - | - | - | - | 1,13 | - |

Chest computed tomography scan showed bilateral ground glass opacities with some areas of consolidation and specimens from his nasopharyngeal swab were positive for COVID-19 on Real Time Reverse-Transcriptase Polymerase Chain Reaction (RT-PCR).

As supplementary oxygen (O2) with a nasal cannula was administered at 2 L/min, his pulse oximetry went up to 97% and he was started on ceftriaxone and clarithromycin. Tacrolimus was suspended and prednisone dose was increased to 30 mg daily.

On day 2 of hospitalization (day 8 of illness) his breathing pattern worsened, as he needed an increase in nasal oxygen flow to 6 L/min, and so did his ascites, which then made his abdomen tense and painful. A diagnostic and therapeutic paracentesis was performed and 2 litres of light yellow ascitic fluid were subsequently removed. Laboratory analysis showed a total cell count of 133 per mm^3^ (macrophages: 60%, mesotelyocytes: 24% and lymphocytes: 16%) and a serum-ascites albumin gradient value of 2. Direct smears and cultures were negative for bacteria, fungi, and mycobacteria, but RT-PCR for SARS-CoV-2 in peritoneal fluid was positive. Serum and fecal samples were also positive (table 2). A rapid serological test performed on day 10 of symptoms was positive for IgM and IgG antibodies.

**Table 2.** Specimen tested for COVID-19 using RT-PCR technique according to date of exam.

| Specimen | 18/05/2020<br>Illness day 5 | 20/05/2020<br>Illness day 7 | 22/05/2020<br>Illness day 9 | 23/05/2020<br>Illness day 10 |
| --- | --- | --- | --- | --- |
| Nasopharyngeal swab | Positive (CT 14-15) | - | - |  |
| Peritoneal fluid | - | Positive (CT 34-38) | - |  |
| Serum | - | - | Positive (CT 31--33) |  |
| Stool | - | - | - | Positive (CT 25-26) |

Sample was considered positive if cycle threshold (CT) value < 40. Lower values suggest higher viral loads.

On day 4, he required intubation as his respiratory status worsened further (PaO2/FiO2 ratio: 110). His ventilation parameters improved in the following days but his renal dysfunction did not and he was started on hemodialysis. The patient died on day 16 due to a refractory septic shock.

## Methods

Nasal and oropharyngeal swabs were collected and stored in 2□mL of sterile Ringer's lactate solution. Ascitic fluid was collected with a sterile syringe following abdominal punction with aseptic technique. Blood was collected in a serum separator tube and then centrifuged according to Centers for Disease Control and Prevention (CDC) guidelines. Stool was collected with a sterile container.

Ribonucleic acid (RNA) extraction was performed performed in all four specimens with QIAamp Viral RNA Mini Kit (QIAGEN, Hilden, Germany), following manufacturer's instructions. Viral detection was performed with AgPath-ID One-Step RT-PCR Reagents (ThermoFisher Scientific, Austin, USA), according to manufacturer’s instructions, in a total of 20 µL reaction volume, containing 5,0 µL of purified RNA, primers and probes (400 nM and 200 nM, respectively), aiming at the N1 and N2 targets of the SARS-CoV-2 nucleoprotein gene, and human ribonuclease P gene as endogenous control, in accordance with CDC guidelines.

Samples with cycle threshold (Ct) values below 40 were considered positive.

This case report was approved by Local Ethics Committee.

## Results

Specimens from nasopharyngeal swab obtained on day 5 of patient’s illness were positive for SARS Cov-2 (table 2). Peritoneal fluid from day 7 was also positive (Ct, 34-38), serum samples collected on day 9 and 10 were positive for both SARS CoV-2 (Ct, 31-32) and IgM/IgG antibodies, respectively. Stool collected on day 10 was positive as well (Ct, 25-26).

## Discussion

A great variety of clinical features have been attributed to COVID-19, including cough and dyspnea^2^, similar to the ones initially reported by this patient. As disease severity progresses, pulmonary injury is found to be a common feature^2^ and previous lung autopsy reports have attributed it to endothelialitis with intracelular viruses^3^. Moreover, as it appears to incite a high rate of thrombotic events^6^, it could also explain further aggravation of a previous portal vein thrombosis observed with this patient, which would justify the worsening of his ascites.

Furthermore, SARS-CoV-2 uses the angiotensin-converting enzyme 2 (ACE2) as a cell receptor to invade human cells^7^ and it is expressed in a wide variety of human tissues, including absorptive enterocytes from ileum and colon^8^, which might have contributed to local viral spread. But more importantly, macrophages also express the ACE2 receptor^9^ and, despite their importance in antiviral defense mechanisms in general, more attention has been given to the hypothesis that, in the case of SARS-CoV-2, they might also enable viral anchoring, replication, and possibly spread to other organs^9^. Since macrophages were predominant in this patient’s ascitic fluid, direct viral injury could also arguably have played a role on initial clinical presentation: an acute viral peritonitis. Interestingly, reports on cirrhotic patients with COVID-19 show a high rate of decompensation with ascites^5^, but this is the first report of SARS CoV-2 RNA detection in ascitic fluid.

It is also important to mention that solid-organ transplant patients on immunosuppressive therapy have impaired T cell immunity, so higher rates of viral replication in general would be expected^10^. On the other hand, SARS-CoV-2 detection in blood samples from immunocompetent patients suggests systemic infection is possible in any case^11^. So, although the combination of a potentially systemic infection in addition to immunosuppression might have contributed to a more severe infection in this patient, the possibility of such clinical scenario in general would imply further investigation in otherwise healthy patients as well, especially due to induced immune dysregulation with T cell exhaustion^4^. Notably, this patient presented with seemingly adequate humoral immune response as he was able to produce IgM and IgG antibodies by day 10 of symptoms onset, which is consonant with previous literature for all groups of patients^12,13^.

Unfortunately, other complications previously described in COVID-19^2^, including renal dysfunction and secondary infection, prolonged hospitalization and led to a fatal outcome.

## Conclusion

This unprecedented case report suggests SARS-CoV-2 can be a direct cause for cirrhosis decompensation and also present as acute peritonitis. More studies are required to better elucidate viral dynamics, immune response and different clinical presentations in distinct populations, especially because of such diverse disease spectrum as seen with COVID-19.

## Data Availability

None.

## Acknowledgements

We thank Dante Cabelho for his assistance with language.

## Authors contributions

Concept, design, administration and writing: VCP, NB

Investigation and interpretation of data: VCP, AHP, LL, DDC, NB

Supervision: OAN, JOA, NB

All authors approve the final version of this manuscript.

